# SARS-CoV-2 Transmission Potential and Policy Changes in South Carolina, February 2020 – January 2021

**DOI:** 10.1101/2021.09.25.21263798

**Authors:** Margaret R. Davies, Xinyi Hua, Terrence D. Jacobs, Gabi I. Wiggill, Po-Ying Lai, Zhanwei Du, Swati DebRoy, Sara Wagner Robb, Gerardo Chowell, Isaac Chun Hai Fung

## Abstract

**Introduction:** We aimed to examine how public health policies influenced the dynamics of COVID-19 time-varying reproductive number (*R*_*t*_) in South Carolina from February 26, 2020 to January 1, 2021.

**Methods:** COVID-19 case series (March 6, 2020 - January 10, 2021) were shifted by 9 days to approximate the infection date. We analyzed the effects of state and county policies on *R*_*t*_ using EpiEstim. We performed linear regression to evaluate if per-capita cumulative case count varies across counties with different population size.

**Results:** *R*_*t*_ shifted from 2-3 in March to <1 during April and May. *R*_*t*_ rose over the summer and stayed between 1.4 and 0.7. The introduction of statewide mask mandates was associated with a decline in *R*_*t*_ (−15.3%; 95% CrI, -13.6%, -16.8%), and school re-opening, an increase by 12.3% (95% CrI, 10.1%, 14.4%). Less densely populated counties had higher attack rate (p<0.0001).

**Conclusion:** The *R*_*t*_ dynamics over time indicated that public health interventions substantially slowed COVID-19 transmission in South Carolina, while their relaxation may have promoted further transmission. Policies encouraging people to stay home, such as closing non-essential businesses, were associated with *R*_*t*_ reduction, while policies that encouraged more movement, such as re-opening schools, were associated with *R*_*t*_ increase.

## Introduction

In late December 2019, a novel virus was reported in Wuhan, China. By January 2020, this virus had been identified as severe acute respiratory syndrome coronavirus 2 (SARS-CoV-2), the causative agent of coronavirus disease 2019 (COVID-19).^1^ The disease was first reported in the United States in January 2020.^2^

With an estimated population of 5,190,705, South Carolina is in the southeastern United States, along the Atlantic coast, and shares borders with North Carolina and Georgia.^3^ South Carolina reported the first case of COVID-19 in the state on March 6, 2020. On March 13, 2020, the Governor of South Carolina declared a State of Emergency.^4^ By April 2, 2020, every county in South Carolina had reported at least one case. Here we report on cases through January 10, 2021, by which point 361,254 cases had been reported, of whom 5,811 died.

Central to infectious disease epidemiology is the concept of the reproduction number (*R*_*0*_) – the average number of secondary cases that a primary case can infect absent any public health intervention in a completely susceptible population.^5^ Before the appearance of the highly transmissible Delta (B.1.617.2) variant, the *R*_*0*_ for COVID-19 was estimated to lie between 2.2,^6^ and 4.4.^7^ In contrast, the time-varying reproduction number (*R*_*t*_) describes the transmission potential at a given timepoint as behavior changes and as public health interventions are implemented.^8^ This makes *R*_*t*_ a better measure of disease spread over time as populations put interventions into effect.^9,10^ Public health policies regarding non-pharmaceutical interventions (NPIs) have been examined for their impact on the *R*_*t*_.^11^ South Carolina put various policies into place from March 16, 2020, through October 5, 2020, primarily in the form of executive orders.

The purpose of this paper is to examine the change in the transmission potential of SARS-CoV-2 in South Carolina over time, especially before and after state or county-level public health policy interventions were put in place. We report the associations of *R*_*t*_ with these policies.

## Methods

This paper uses historic COVID-19 data from March 6, 2020 – January 10, 2021 from all 46 counties of South Carolina. South Carolina’s Department of Health and Environmental Control divides the state into four regions: Upstate, Midlands, Pee Dee, and Low Country. A map of all counties in South Carolina by health region is provided in **Supplemental Figure 1**. Population by county is presented in **Supplemental Figure 2**. Cumulative case count and cumulative case count per 100,000 population, in April, August, October and December 2020 are presented in **Supplemental Figure 3**.

Information about policies enacted in South Carolina was obtained from Executive Orders published online by the government of South Carolina. County level policies were obtained from county health departments. Information about school openings was obtained from school district websites, and in cases where schools had staggered openings (for example middle schools starting before high schools), the earlier date was used. Detailed information on these policies including the date of the implementation and relaxation of public health interventions is presented in **Table 1**.

**Table 1.**
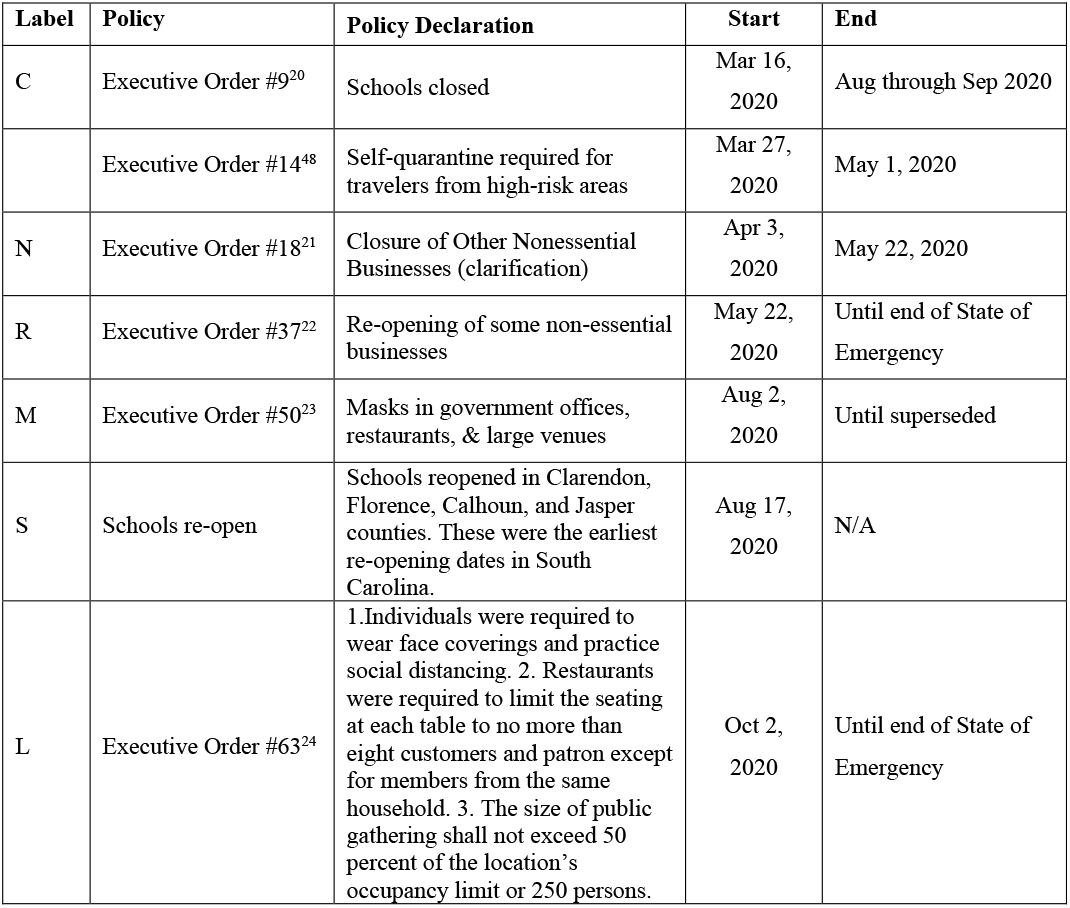
Policies enacted in South Carolina either by Executive Order or by local school districts, in the case of school re-opening. The labels correspond to Figures 1, 2, and 3 where appropriate.

| Label | Policy | Policy Declaration | Start | End |
| --- | --- | --- | --- | --- |
| C | Executive Order #9<sup>20</sup> | Schools closed | Mar 16, 2020 | Aug through Sep 2020 |
|  | Executive Order #14<sup>48</sup> | Self-quarantine required for travelers from high-risk areas | Mar 27, 2020 | May 1, 2020 |
| N | Executive Order #18<sup>21</sup> | Closure of Other Nonessential Businesses (clarification) | Apr 3, 2020 | May 22, 2020 |
| R | Executive Order #37<sup>22</sup> | Re-opening of some non-essential businesses | May 22, 2020 | Until end of State of Emergency |
| M | Executive Order #50<sup>23</sup> | Masks in government offices, restaurants, & large venues | Aug 2, 2020 | Until superseded |
| S | Schools re-open | Schools reopened in Clarendon, Florence, Calhoun, and Jasper counties. These were the earliest re-opening dates in South Carolina. | Aug 17, 2020 | N/A |
| L | Executive Order #63<sup>24</sup> | 1. Individuals were required to wear face coverings and practice social distancing. 2. Restaurants were required to limit the seating at each table to no more than eight customers and patron except for members from the same household. 3. The size of public gathering shall not exceed 50 percent of the location's occupancy limit or 250 persons. | Oct 2, 2020 | Until end of State of Emergency |

### Data acquisition

From the New York Times GitHub repository,^12^ we downloaded the cumulative confirmed COVID-19 case count from March 6, 2020 - January 10, 2021. Data for the daily incidence were cleaned at the county level to eliminate any dates with negative incidence (**Appendix A**). We selected a starting point of March 6, 2020 since the first case in South Carolina was reported on this date and a cutoff date of January 10, 2021 for all analyses. A nine-day backward shift was used to estimate the date of infection, accounting for a 6-day incubation period and a 3-day delay in testing. The error of this simple approach is considered tolerable if the delay to observation is not highly variable and if the mean delay is known.^8^ This translated into the assumed date of infection from February 26, 2020 to January 1, 2021. Our choice of the cut-off point allowed us to complete the time series by the end of winter holiday season (Christmas to New Year). A sensitivity analysis was conducted using a lower boundary of 4 days and an upper boundary of 15 days based on incubation data reported by McAloon et al.,^13^ and CDC reports on testing delays.^14^ We assessed the 2019 county-level population data for South Carolina from the U.S. Census Bureau^15^ and examined the power-law relationship between cumulative case count and population size through linear regression between the log_10_-transformed per capita cumulative case number and log_10_-transformed population size for each county.

### Statistical Analysis

To calculate *R*_*t*_, we used the instantaneous reproduction number method in the R package *EpiEstim* with the parametric option. This measure of the *R*_*t*_ was defined by Cori et al.^16^ as the ratio between *I*_*t*_, the number of incident cases at the time *t*, and the total infectiousness of all infected individuals at the time *t*. This *R*_*t*_ was used to describe the burden of COVID-19 at a state level and throughout certain counties.

The *R*_*t*_ is presented in two ways in this paper. The first way is to take the average of the daily *R*_*t*_ estimates over a 7-day sliding window. The second way is to take the average of the daily *R*_*t*_ estimates over a non-overlapping time window between two time points of policy changes (hereafter, known as policy change *R*_*t*_ in this paper).

The policy change *R*_*t*_ was used to establish associations with policies. We calculated the percentage change for the policy change *R*_*t*_ for South Carolina (**Supplemental Table 1**), using the median policy change *R*_*t*_ estimate at each policy interval. For instance, the median policy change *R*_*t*_ estimate at each policy interval will be compared to the previous policy interval, as in 100%×(t_2_-t_1_)/t_1_. We utilized EpiEstim *sample from the posterior R distribution* function to obtain a sample of 1000 estimates of *R*_*t*_ for each t_1_ and t_2_ then estimate the credible intervals (2.5 and 97.5 percentile) of the percentage change. We also calculated the percentage change of the policy change *R*_*t*_ for Beaufort, Calhoun, Charleston, Colleton, Georgetown, Oconee, Orangeburg, Richland, and Williamsburg counties in South Carolina (**Supplemental Table 2**). These nine counties were selected because they are the only counties with an active mask ordinance during the study period.

We characterized the power-law relationship between the county-level cumulative number of COVID-19 cases and population size, following C∼N^g (C, cumulative case count; N, population size; g, exponent).^17^ We performed linear regression analysis between the log_10_-transformed per capita cumulative case count and the log_10_-transformed population size, i.e., log_10_(C/N) = m log_10_(N) where m = g-1.^17-19^ We computed linear regression between the log_10_-transformed per capita cumulative case count and the log_10_-transformed population size, at four different dates: June 30^th^, August 31^st^, October 31^st^, and December 31^st^.

When per capita cumulative case count is proportional to population size, then there was no heterogeneity of per capita cumulative case count across geographic units (counties) of different population sizes (i.e., when m=0 and thus g=1). Geographical units with lower population sizes would have a higher per capita cumulative case count if m<0 (i.e., g<1) and those with lower population sizes would have a lower per capita cumulative case count if m>0 (i.e., g>1).^18,19^ See **Appendix B** for details.

Statistical analysis was performed using R 4.0.3 (R Core Team, R Foundation for Statistical Computing, Vienna, Austria). **Supplemental Figures 1, 2 and 3** were created using R 3.5.1 (R Core Team, R Foundation for Statistical Computing, Vienna, Austria).

### Ethics

The Georgia Southern University Institutional Review Board made a non-human subject determination for this project (H20364) under the G8 exemption category according to the Code of Federal Regulations Title 45 Part 46.

## Results

### State Level - General

The daily new case count rose at the beginning of June 2020, and the peak of the first wave of cases arrived by mid-July. Case counts then started falling but remained higher than the beginning of the pandemic. By late September, case count rose again, and continued to rise through the end of the study period. Several days were reported with 0 cases, as data was not reported on federal holidays (Thanksgiving, Christmas Day, and New Year’s Day). **Figure 1** displays daily incident case count, 7-day sliding window *R*_*t*_, and the policy change *R*_*t*_, all right-adjusted for nine days at the state level. The 7-day sliding window *R*_*t*_ throughout the state fluctuated between 2 and 3 in early March, and decreased to <1 during parts of April and May 2020. Over the summer, the *R*_*t*_ rose and continued to fluctuate between 0.7 and 1.4 throughout the state. At the end of the study period, the *R*_*t*_ was still above 1.0 indicating continued spread of the virus.

**Figure 1.**
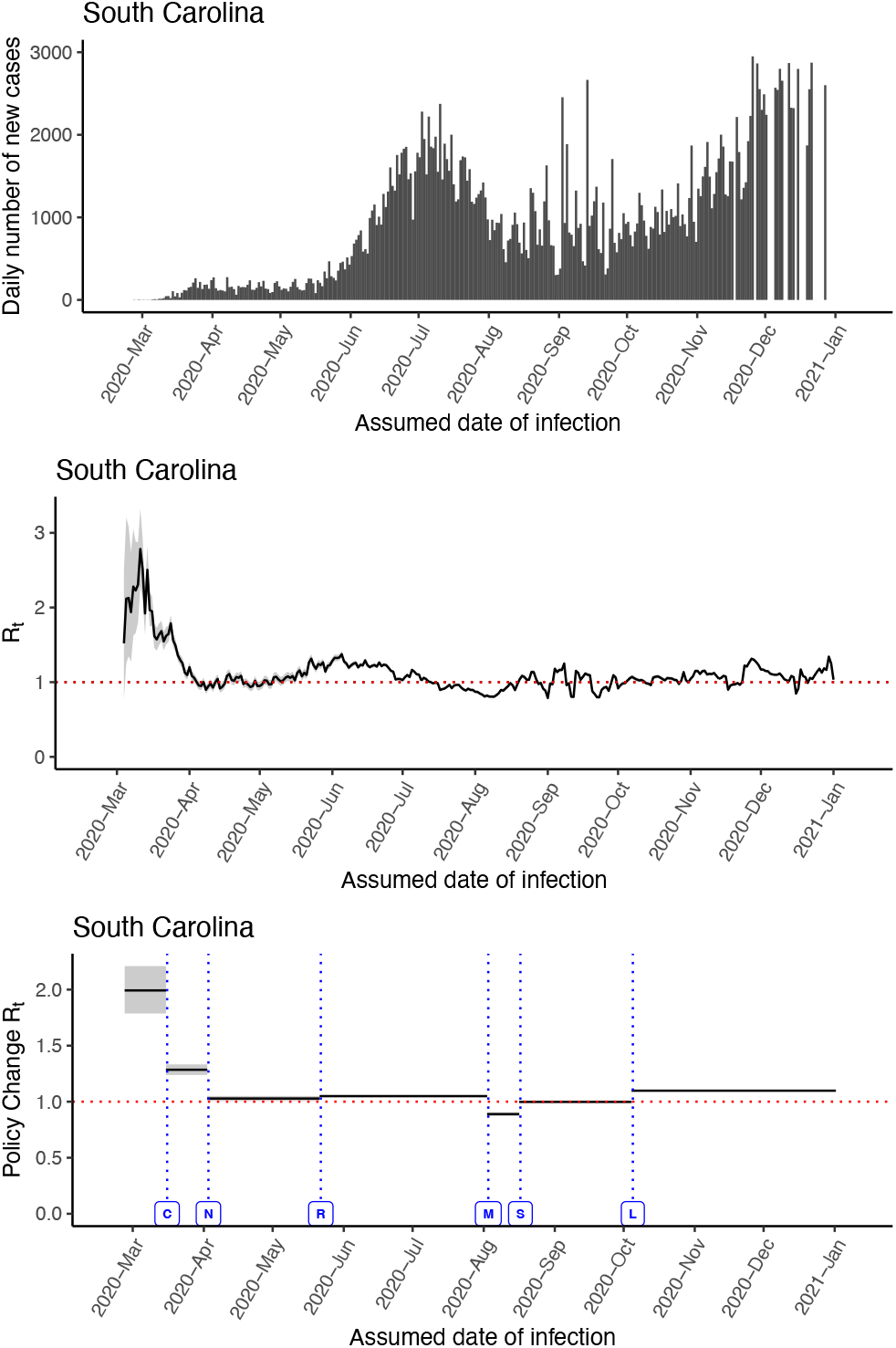
Daily number of new cases, 7-day sliding window *R*_*t*_, and Policy Change *R*_*t*_ for the state of South Carolina. All case count data have been shifted backwards by 9 days to approximate the date of infection. Data were not reported on holidays (Thanksgiving, Christmas, and New Year’s Day). Policy Change *R*_*t*_ labels: C: Closure of schools (Mar 16, 2020), N: Closure of non-essential businesses (Apr 3, 2020), R: Re-opening of non-essential businesses (May 22, 2020), M: State level mask mandate (Aug 2, 2020), S: Re-opening of schools using earliest reported date (Aug 17, 2020), L: Capacity limits on public gathering changed to 250 (Oct 2, 2020).

### Policy Impacts – State Level

*R*_*t*_ fluctuated with policy changes at the state level. The *R*_*t*_ presented in this section refers to the policy change *R*_*t*_. (**Figure 1**: lower panel, **Supplemental Table 1**). Prior to the introduction of any policies, the *R*_*t*_ was 1.991 (95% credible interval, CrI, 1.787, 2.21). The first policy introduced was the closure of schools on March 16^th^.^20^ Between the closure of schools and the closure of non-essential businesses, the *R*_*t*_ was 1.285 (95% CrI, 1.24, 1.33), a decrease of 35.59% (95% CrI, 27.9%, 42.7%).

The closure of non-essential businesses was ordered on April 3,^21^ indicated by the label ‘N’ in **Figure 1**. *R*_*t*_ dropped to 1.028 (95% CrI, 1.01, 1.05), a decrease of 20.01% (95% CrI, 18.8%, 21.1%), although the policy window was short. Some non-essential businesses were allowed to begin re-opening on May 22, following the issue of Executive Order 37.^22^ The *R*_*t*_ associated with this timeframe was 1.05 (95% CrI, 1.04, 1.06), a statistically insignificant increase of 2.07% (95% CrI, -0.217 %, 4.2%).

The next Executive Order we examined was passed on August 2, 2020 mandating masks in government building, restaurants, and large venues.^23^ This was associated with the first occurrence of *R*_*t*_ dropping below 1.0 in our policy examination. During this timeframe, the *R*_*t*_ was 0.889 (95% CrI, 0.873, 0.905), a decrease of 15.3% (95% CrI, 13.6%, 16.8%).

Our proxy date for school openings was August 17, 2020. This was based on the earliest reported dates for school openings. The *R*_*t*_ rose following this date to 0.998 (95% CrI, 0.989, 1.01), an increase of 12.3% (95% CrI, 10.1%, 14.4%). The final policy in this analysis was enacted on October 2, allowing restaurants to reopen for indoor dining and lifting capacity limits.^24^ This was followed by an increase in *R*_*t*_ to 1.098 (95% CrI, 1.09, 1.1), increasing by 9.994% (95% CrI, 9.47%, 10.5%). This indicated sustained transmission of COVID-19 in South Carolina.

Sensitivity analysis was conducted to examine the effect of the assumption of the time lag. A 15-day time lag (**Supplemental Figure 4**) and a 4-day time lag (**Supplemental Figure 5**) were applied to the time series of the state-level case count data and no major differences between the main results and the lagged results were observed.

### Mask Mandates – County Level

The wearing of masks has been advised for the general public since early April of 2020.^25^ However, the requirement for face mask wearing was left up to each state, likely due to the federal polity of the U.S. and the political atmosphere in 2020.^26^ For the purposes of this paper a “mask mandate” is any order given by authority for residents of a certain area to wear a mask or face covering while in specified locations. In South Carolina, the first Executive Order to mandate masks was issued on August 3, 2020.^20^ This Order only mandated masks in government buildings, restaurants, and large venues.

Several counties (Beaufort, Charleston, Georgetown, Orangeburg, Richland, and Williamsburg) issued their own mask mandates before the state. Three counties (Calhoun, Colleton, and Oconee) issued a mask mandate after the statewide order was passed. We showed the policy change *R*_*t*_ for these counties in **Figure 2** and **Figure 3**. These nine counties were the only counties with an active mask ordinance during the study period (**Supplemental Table 2**).

**Figure 2.**
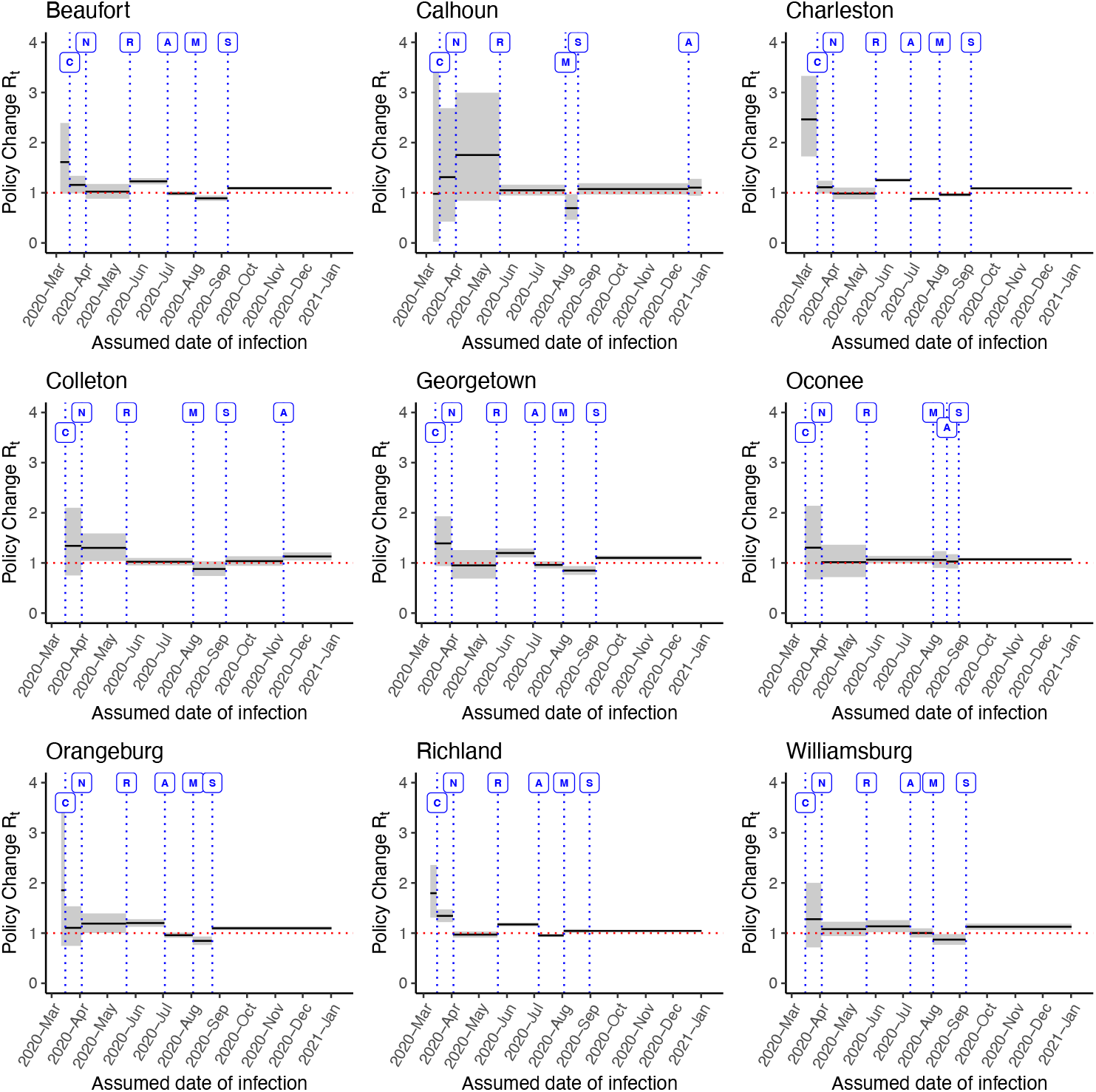
Policy change *R*_*t*_ in counties with mask mandates in South Carolina. Labels – C: Closure of schools (Mar 16, 2020), N: Closure of non-essential businesses (Apr 3, 2020), R: Re-opening of nonessential businesses (May 22, 2020), A: County level mask mandate (Jul 3, 2020), M: State mask mandate (Aug 2, 2020), S: Start of school, based on the earliest date in the county (Sep 8, 2020). County locations can be found in Supplemental Figure 1.

**Figure 3.**
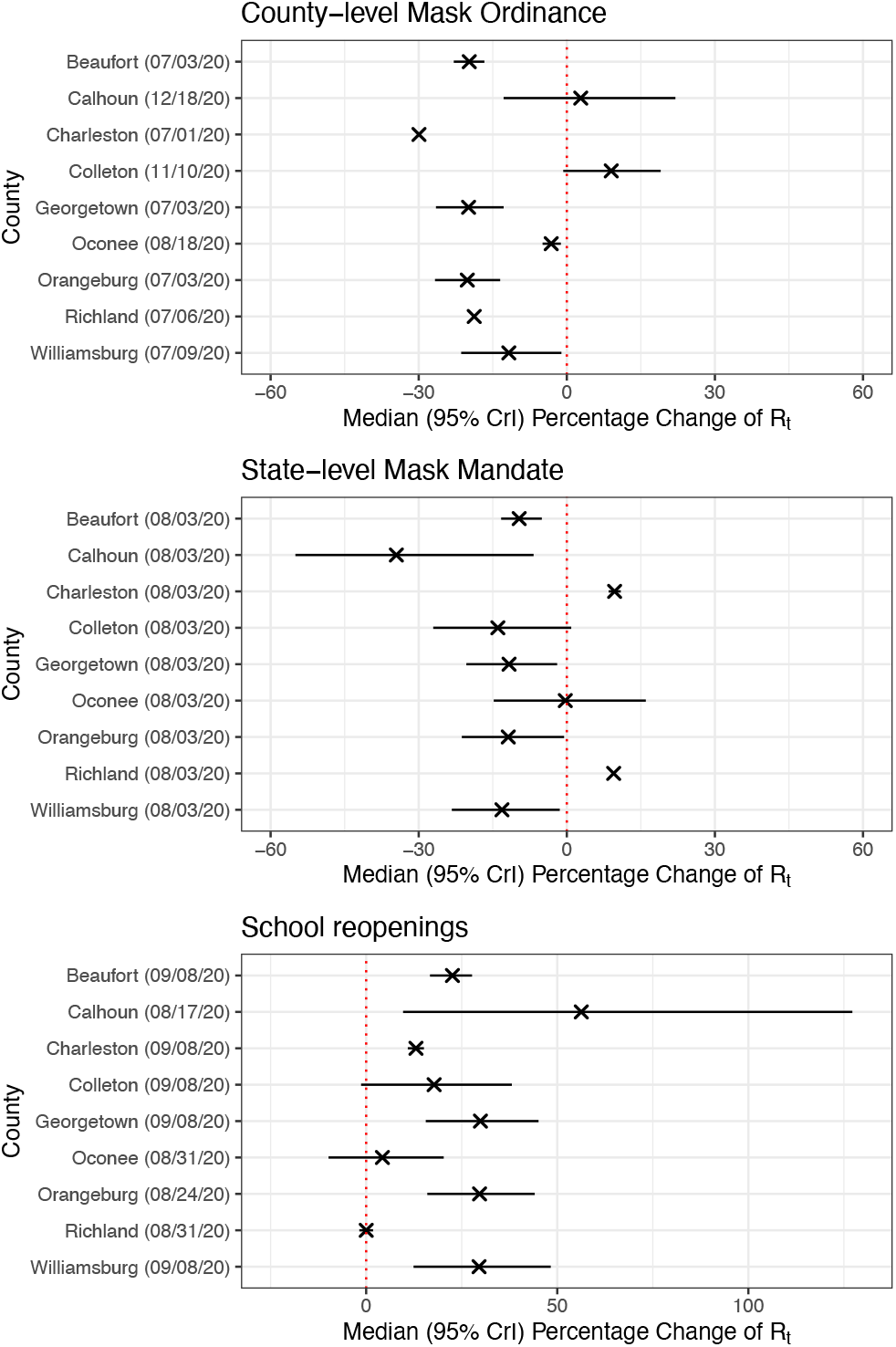
Median percentage change (95% CrI) of policy change *R*_*t*_ estimates for nine selected counties organized by non-pharmaceutical interventions (county-level mask ordinance, state-level mask mandate, and school re-openings). The vertical line at zero percentage change on the x-axis indicated an increase in *R*_*t*_ (positive percentage changes) to its right and a decrease in *R*_*t*_ (negative percentage changes) to its left.

The first counties we examined were those that passed the county-level mask ordinance before the state mandate. Beaufort county passed its ordinance on July 3^th^ 2020.^27^ The *R*_*t*_ decreased from 1.2283 (95% CrI, 1.17, 1.29) to 0.9856 (95% CrI, 0.946, 1.027), a decrease of 19.76% (95% CrI, 16.7%, 22.9%). Charleston county passed its first mask ordinance on July 1^st^, 2020.^28^ After the county ordinance passed, the *R*_*t*_ dropped from 1.2526 (95% CrI, 1.22, 1.28) to 0.8774 (95% CrI, 0.855, 0.90), decreasing by 29.95% (95% CrI, 29.9%, 30.0%). Georgetown County’s mask ordinance passed on July 3^rd^, 2020.^29^ The *R*_*t*_ decreased to 0.9596 (95% CrI, 0.891, 1.032) from 1.1980 (95% CrI, 1.11, 1.29), a decrease of 19.89% (95% CrI, 12.8%, 26.5%). Orangeburg County passed its Face Mask Ordinance on July 3^rd^, 2020,^30^ and its *R*_*t*_ estimates decreased from 1.2002 (95% CrI, 1.13, 1.28) to 0.9585 (95% CrI, 0.908, 1.011) with a decrease of 20.16% (95% CrI, 13.5%, 26.7%). Richland County passed its Face Mask Ordinance on July 6^th^, 2020,^31^ and its *R*_*t*_ estimates decreased from 1.1729 (95% CrI, 1.14, 1.21) to 0.9529 (95% CrI, 0.922, 0.984), a decrease of 18.76% (95% CrI, 18.7%, 18.8%). Finally, Williamsburg County’s *R*_*t*_ decreased after the introduction of their mask mandate on July 9^th^, 2020,^32^ from 1.1342 (95% CrI, 1.02, 1.26) to 1.0013 (95% CrI, 0.912, 1.069), a decrease of 11.75% (95% CrI, 1.1%, 21.4%).

Calhoun, Colleton, and Oconee counties had their county level mask mandates passed after the August 3^rd^ state mask mandate (**Figure 3**, top panel). Among them, Oconee County passed a county-level mask ordinances on August 18^th^ before the school re-opening.^33^ After the county-level mandate passed, the *R*_*t*_ further decreased from 1.0591 (95% CrI, 0.901, 1.235) to 1.0259 (95% CrI, 0.89, 1.17), a decrease of 3.16% (95% CrI, 1.21%, 4.91%). However, Calhoun County and Colleton County passed their county-level face mask ordinances much later than the state level mask mandate and months after the schools reopened in the Fall. Colleton County passed the county-level face mask ordinances on November 10^th^, 2020,^34^ and the *R*_*t*_ increased by 9.0% (95% CrI: -0.719%, 19.01%) from 1.0353 (95% CrI: 0.943, 1.133) to 1.1287 (95% CrI: 1.05, 1.21), but the increase was statistically insignificant. Lastly Calhoun County passed their county-level face mask ordinances on December 18^th^, 2020,^35^ and the *R*_*t*_ increased from 1.0737 (95% CrI: 0.965, 1.19) to 1.101 (95% CrI: 0.941, 1.278), but the increase of 2.82% (95% CrI, - 12.8%, 22.0%) was statistically insignificant.

### School Openings

School openings were examined in both **Figures 1, 2** and **3**. In **Figure 1**, school re-opening is indicted by label ‘S’, where we used a proxy date of August 17^th^, the earliest reported school opening date across the state. It is important to note that these school openings are based on K through 12 schools’ starting dates and not college starting dates. Some schools reopened in a staggered way by grade. Following school openings, the *R*_*t*_ in South Carolina rose by 12.3% (95% CrI, 10.1%, 14.4%) from 0.889 (95% CrI, 0.873, 0.91) to 0.998 (95% CrI, 0.989, 1.01).

At the county level (**Figures 2 and 3**), *R*_*t*_ increased when schools were re-opened in most counties. In Beaufort County, the increase was 22.57% (95% CrI, 16.7%, 27.7%) from 0.8903 (95% CrI, 0.839, 0.944) to 1.0916 (95% CrI, 1.06, 1.12). Calhoun County’s *R*_*t*_ increased from 0.6857 (95% CrI, 0.461, 0.974) to 1.0737 (95% CrI, 0.965, 1.19), an increase of 56.29% (95% CrI, 9.66%, 127.21%). In Charleston County, the *R*_*t*_ rose by 13.03% (95% CrI, 10.9%, 15.1%) from 0.9621 (95% CrI, 0.927, 0.998) to 1.0878 (95% CrI, 1.07, 1.11). The Colleton County *R*_*t*_ increased by 17.77% (95% CrI, -1.33%, 38.12%) from 0.8782 (95% CrI, 0.74, 1.03) to 1.0353 (95% CrI, 0.943, 1.133), but the increase was statistically insignificant. The *R*_*t*_ in Georgetown County rose from 0.8468 (95% CrI, 0.762, 0.937) to 1.1016 (95% CrI, 1.06, 1.14), increasing by 29.9% (95% CrI, 15.6%, 45.1%).

Oconee County had a statistically insignificant increase of 4.25% (95% CrI, -9.85%, 20.29%) in *R*_*t*_ from 1.0259 (95% CrI, 0.89, 1.17) to 1.0710 (95% CrI, 1.04, 1.11). Orangeburg County had an increase in *R*_*t*_ from 0.8447 (95% CrI, 0.763, 0.932) to 1.0972 (95% CrI, 1.06, 1.14), an increase of 29.67% (95% CrI, 16.0%, 44.1%). In Richland County, the *R*_*t*_ increased slightly from 1.0434 (95% CrI, 1.01, 1.08) to 1.0439 (95% CrI, 1.03, 1.06), but the increase of 0.016% (95% CrI, -1.78%, 1.79%) was statistically insignificant. Williamsburg County’s *R*_*t*_ increased to 1.1264 (95% CrI: 1.07, 1.19) from 0.8692 (95% CrI: 0.767, 0.98), an increase of 29.54% (95% CrI: 12.4%, 48.3%).

### Power-law Relationship between Cumulative Case Number and Population Size

**Figure 4** presents the linear regression models between the log_10_-transformed per capita cumulative case number and the log_10_-transformed population size for a total of 46 counties in South Carolina at four different dates of report, June 30^th^, August 31^st^, October 31^st^, and December 31^st^, 2020, respectively. Each regression line represents a specific assessed date (date of report); and the slopes, m, of four regression lines were calculated and documented in **Table 2**. Slopes of four regression lines were negative (m<0) and statistically significant (m = -2.0236, -1.2164, -1.0220, -1.0577; p<0.0001 respectively). A negative slope suggests that counties with lower population sizes (i.e., rural counties) would have a higher per capita cumulative case count. This result suggests the existence of potential health disparities between urban and rural counties.

**Table 2.** The slope (and 95% Confidence Intervals) of the regression line between log_10_-transformed per capita cumulative count and log_10_-transformed population size by county in South Carolina, USA, on June 30th, August 31st, October 31st, and December 31st, 2020 (date of report).

| Date of case report | Slope (m) and 95% CI | P-value |
| --- | --- | --- |
| June 30th, 2020 | -2.0236 (-2.8233, -1.2239) | p<0.0001 |
| August 31st, 2020 | -1.2164 (-1.6712, -0.7615) | p<0.0001 |
| October 31st, 2020 | -1.0220 (-1.4088, -0.6352) | p<0.0001 |
| December 31st, 2020 | -1.0577 (-1.4456, -0.6697) | p<0.0001 |

**Figure 4.**
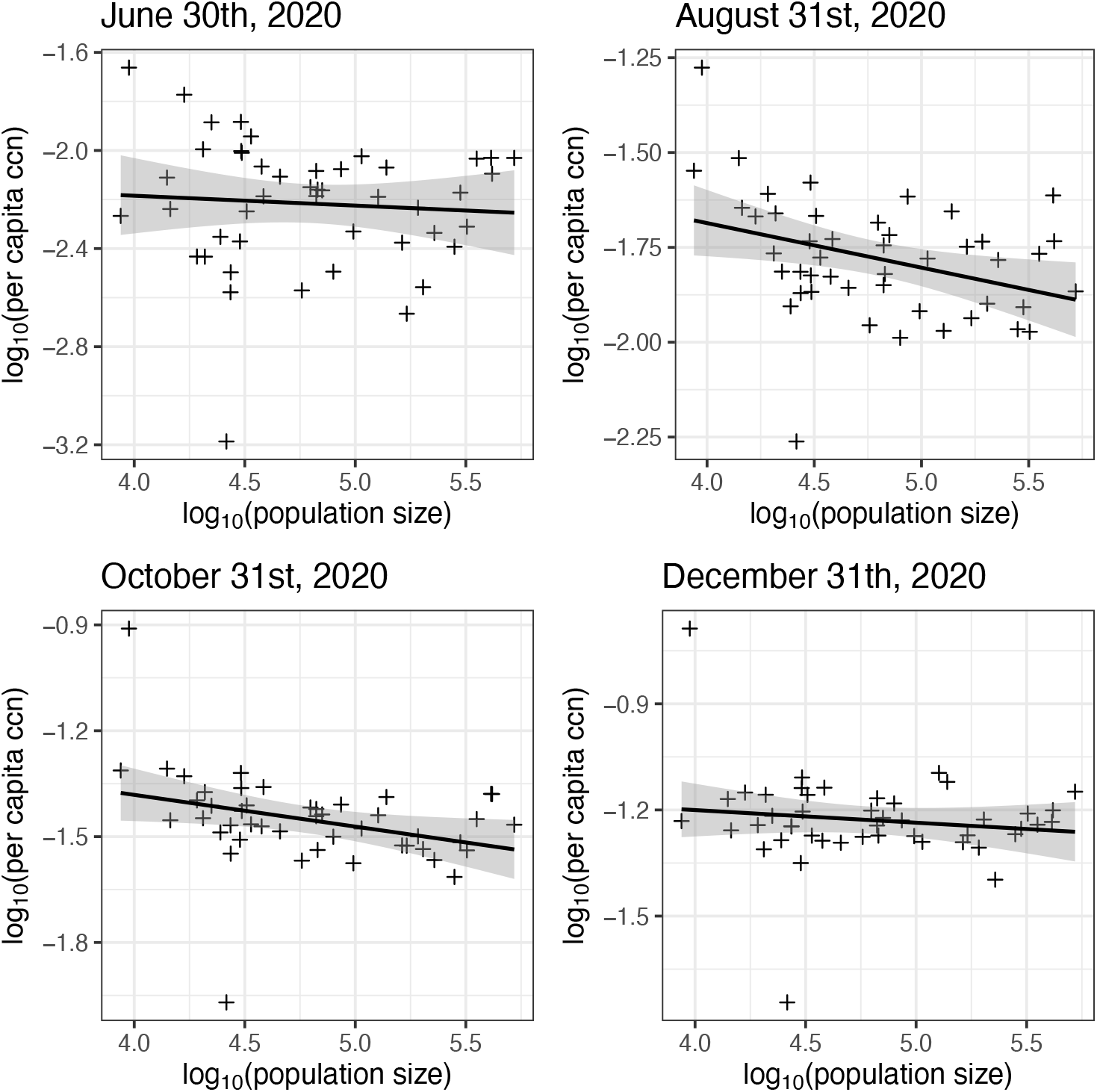
Linear regression between log_10_-transformed per capita cumulative case number (ccn) and log_10_-transformed population size by county for South Carolina on June 30th, August 31st, October 31st, and December 31st, 2020. Each plus sign represents a county in South Carolina.

## Discussion

This paper examined the associations between SARS-CoV-2 *R*_*t*_ and public health policy changes throughout South Carolina from February 2020 to January 2021. We specifically examined the impacts of mask mandates at a county level and the resumption of in-person school activities (**Figures 2 and 3**). We found that mask mandates were frequently associated with a decrease in *R*_*t*_ while school re-openings were frequently associated with an increase in *R*_*t*_.

We found that in Beaufort, Charleston, Georgetown, Oconee, Orangeburg, Richland, and Williamsburg counties, where a mask mandate was introduced at the county level prior to the state level mandate, a decrease in *R*_*t*_ was associated with the introduction of the policy. This suggested that county-level facemask mandate did have its utility in dampening SARS-CoV-2 transmission. In Oconee County, the state-level facemask mandate happened before the county-level facemask mandate. The county-level mandate apparently led to a slight further decrease in *R*_*t*_.

In two counties, Calhoun and Colleton, the introduction of a county-level mask mandate happened late in 2020, after the introduction of the state-level mask mandate and the re-opening of schools. In both cases, *R*_*t*_ dropped below 1 after the state mask mandate but increased to levels above 1 after schools reopened. Our results suggest that the county-level mandates were introduced too late to have a significant impact on *R*_*t*_. The increased *R*_*t*_ after the county-level mask mandate should be interpreted as a continuation of an increase in *R*_*t*_ despite the county-level mask mandate. Additionally, by late 2020 adherence fatigue^36^ might also impact how well facemask mandates were followed.

We also examined school re-openings in counties that had county level mask mandates in place (**Figures 2 and 3**). In these counties, the *R*_*t*_ fell when the county mask mandate was put in place. In most places the *R*_*t*_ lowered again when the state mandate was put into place, although in Charleston and Richland counties the *R*_*t*_ did rise after the state mandate. This may be due to adherence fatigue^37^ in the summer months. Richland county includes Columbia, the state capital. Columbia is highly populated and is the site of the University of South Carolina main campus. Case counts here might be impacted by the university opening (such as student parties that turned out to be super-spreading events).^38^ Charleston is a tourist destination, so potentially an increase in late summer tourism could have driven the *R*_*t*_ higher despite the statewide mask mandate, especially as the mandate only required masks in government buildings, restaurants, or large venues. Similar observations can be said of Beaufort County, where the tourist destination Hilton Head Island is located. While the county’s and state’s mask mandates were associated with *R*_*t*_ decreasing to below 1 in Beaufort, *R*_*t*_ increased after school re-opening in September.

Other literature supports the notion that mask mandates may slow the transmission of SARS-CoV-2.^39^ Hua et al. found that a mask mandate was associated with a decrease in *R*_*t*_ by 27% in North Dakota, by 16% in Montana and by 13% in Wyoming.^40^ Politis et al. found facemask mandate was associated with a decrease in *R*_*t*_ by 11% and 6%, respectively, in Arkansas and Kentucky.^10^ Thus, our findings in South Carolina are consistent with findings in other states that a mask mandate was associated with slowing epidemic growth.

The role of school re-openings in COVID-19 transmission has been examined as well.^37,41-44^ A high school in Israel reported a COVID-19 outbreak shortly after a school reopened in May 2020.^42^ Another study modeled school re-opening, and found that reduction in capacity and mask wearing could reduce community transmission, whereas higher capacity and non-adherence to mask wearing could drive COVID-19 spread in the school’s community.^44^ According to our analyses (**Figures 2 and 3**), the percentage changes of policy change *R*_*t*_ estimates increased in eight of nine selected counties, except Richland County. This observation echoes existing studies that school re-openings have the potential to spread COVID-19 in the local communities.^37,42-44^ Similar to our findings, Hua et al. found an increase in *R*_*t*_ in Idaho (13%), Montana (21%), South Dakota (12%) and Wyoming (20%) after school reopened on September 7, 2020; however, the same study found a decrease in *R*_*t*_ by 8% in North Dakota after school re-opening on the same date.^40^ Politis et al. found that after school reopened, *R*_*t*_ increased by 12% and 9% in Arkansas and Kentucky, respectively.^10^ Thus, our findings in South Carolina are consistent with findings in other states in general, that school re-opening in August and September 2020 was associated with increased SARS-CoV-2 transmission as evidenced in an increase in *R*_*t*_.

In addition, rural counties in South Carolina were found to have a higher per capita cumulative case count at four different assessed dates in 2020. This result suggests that rural counties bore a higher disease burden than urban counties. Future research may investigate the cause and related factors of such health disparities.

The focus of this study was on public health and social policy involving mandates of NPIs. Our study period ended in early 2021 before the vaccination campaign could make an impact to slow SARS-CoV-2 transmission in South Carolina. Future research may study whether certain highly transmissible variants of concern may trigger COVID-19 resurgence.^45^ While this is out of scope for this study, further research into the effect of policy mandates that target special populations such as residents of long-term care facilities and their caretakers may be conducted.^46^

## Limitations

There are a number of limitations in this study. First, this analysis was based on aggregate data reported by the surveillance system of COVID-19 in South Carolina. The data was arranged by date of report. Even though we shifted the date backward by 9 days to approximate the date of infection, this remained an estimation. Second, date of report is affected by holidays, on which days cases were not reported. Third, the effects of viral variants on transmission potential^45^ cannot be shown in this study. The first two cases of the Beta (B.1.351) variant in the U.S. were detected in South Carolina after the study period ended,^47^ so this may not be a severe limitation. Fourth, while re-opening of schools was staggered by grade in South Carolina, we lumped the re-openings together as we chose the first date of the re-opening as the date of policy change. However, for the county-level policy change analysis, we had specific school re-opening dates for all nine selected counties (**Figures 2 and 3**). And finally, we do not examine the impact of vaccinations on the transmission potential in South Carolina; however, our study period ended by January 10^th^, 2021 (date of report), by which point there were minimal numbers of people fully vaccinated.

Although we examined the impact of policy mandates, we did not examine the extent to which these policies were adhered on the ground. Behavioral variation in some places might impact the effectiveness of policies. However, as we attempted to examine the real-world effects of interventions on COVID-19 transmission potential, this would not be a serious limitation to the paper.

## Conclusions

The pandemic affected South Carolina starting with the first cases confirmed in early March 2020, and data suggest ongoing transmission from late February 2020 through the end of the study period (the beginning of 2021). Our findings suggest that public health policies that encourage the adoption of NPIs, such as mask mandates, were found to be associated with a decrease in *R*_*t*_. In contrast, policies that encouraged more social interaction and population movement, such as re-opening schools for in-person instruction, were typically followed by an increase in *R*_*t*_. In general, mask mandates appeared to work better in counties that implemented it early on than those that implemented it after the incidence trajectory had risen to a high level. Our paper provided a state and county-level analysis that could support evidence-based decision-making in the adoption of NPIs at the population level against COVID-19. Our findings could prove useful for shaping future outbreak responses.

## Data Availability

The data analyzed in this manuscript are publicly available at the New York Times GitHub repository (https://github.com/nytimes/covid-19-data).

## Acknowledgement

The authors thank Prof. Anne C. Spaulding for her review of an earlier draft of the manuscript and for her helpful suggestions.

## List of abbreviations

COVID-19: Coronavirus disease 2019
NPIs: Non-Pharmaceutical Interventions
*R*_*0*_: Basic reproduction number
*R*_*t*_: Time-varying reproduction number
SARS-CoV-2: Severe acute respiratory syndrome coronavirus 2
SoE: State of Emergency

## Supplemental Materials

## Appendix A: Handling of negative incident case count

A negative incident case count means the daily number of new cases reported being negative. This happened when the local public health department made an adjustment to the previously reported data by removing the duplicated cases or cases introduced by human errors. This usually resulted in a cumulative case count of the day being lower than the cumulative case count of the previous day, which translates into a negative incident case count. When negative cases appeared in the downloaded data, we used the previous day’s case data to bring the negative incidence to zero. In the instance when the previous day did not have enough cases to make the negative incidence zero, we worked backward until there were enough cases brought to the negative case count to equal it to zero.

## Appendix B: Cumulative case count and Population size of a County

The power-law relationship between the county-level cumulative number of COVID-19 cases and population size can be transformed into a linear relationship between the log_10_-transformed cumulative case count and the log_10_-transformed population size as follows^1,2^:

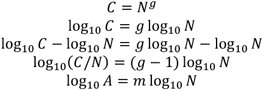

where the per capita cumulative case count A=C/N and the slope of the regression line, m = g-1.

In this paper, we performed linear regression models between log_10_-transformed per capita cumulative case count and log_10_-transformed population size of counties in South Carolina. The data analyzed were by the dates of report of June 30, August 31, October 31 and December 31, 2020. If the slope *m* is positive, it implies that counties with higher populations (i.e., urban counties) had a higher attack rate. If the slope is negative, it implied that counties with lower populations (i.e., rural counties) had a higher attack rate. If the slope is 0 (or if the 95% confidence interval includes 0), it implied that the attack rate was the same across the counties regardless of their population size.

---

**Supplemental Table 1.** Difference in Policy Change *R*_*t*_ as policies changed at South Carolina (state level).

| State | Median $R_t$ & 95%CrI | Median $R_t$ difference percentage changes comparing with previous policy interval & 95%CrI |
| --- | --- | --- |
| Before policy C 1 | 1.991 (1.787, 2.21) |  |
| C→ N 1,2 | 1.285 (1.24, 1.33) | -35.59% (-27.9%, -42.7%) |
| N→ R 2,3 | 1.028 (1.01, 1.05) | -20.01% (-18.8% -21.2%) |
| R→ M 3,4 | 1.05 (1.04, 1.06) | 2.07% (-0.217%, 4.2%) |
| M→ S 4,5 | 0.889 (0.873, 0.905) | -15.3% (-13.6%, -16.8%) |
| S→ L 5,6 | 0.998 (0.989, 1.01) | 12.3% (10.1%, 14.4%) |
| Beyond L 6+ | 1.098 (1.09, 1.1) | 9.994% (9.47%, 10.5%) |

CrI, credible intervals. Policy Change $R_t$ labels: C: Closure of schools, N: Closure of non-essential businesses, R: Reopening of non-essential businesses, M: state level mask mandate, S: Re-opening of schools (earliest reported date), L: Capacity limits on public gathering changed to 250.

**Supplemental Table 2.** Difference in Policy Change *R*_*t*_ as policies changed at county levels in selected counties South Carolina. These nine counties were the only counties with an active mask ordinance during the study period.

| | Median $R_t$ & 95%CrI | Median $R_t$ difference percentage changes comparing with previous policy interval & 95%CrI |
| --- | --- | --- |
| **Beaufort** |  |  |
| Before policy C | 1.5853 (0.985, 2.392) |  |
| C → N | 1.1551 (0.99, 1.34) | -26.74% (-54.6%, 16.9%) |
| N → R | 1.0214 (0.881, 1.175) | -11.58% (-11.0%, -12.1%) |
| R → A | 1.2282 (1.17, 1.29) | 20.04% (2.54%, 40.03%) |
| A → M | 0.9856 (0.946, 1.027) | -19.76% (-16.7%, -22.9%) |
| M → S | 0.8903 (0.839, 0.944) | -9.65% (-5.07%, -13.29%) |
| S → | 1.0916 (1.06, 1.12) | 22.57% (16.7%, 27.7%) |
| **Calhoun** |  |  |
| Before policy C | 0.6787 (0.0248, 3.6122) |  |
| C → N | 1.2254 (0.426, 2.687) | 82.83% (-74.7%, 3840.2%) |
| N → R | 1.6951 (0.841, 2.995) | 39.55% (-50.2%, 331.3%) |
| R → M | 1.0495 (0.945, 1.161) | -38.51% (-66.4%, 24.1%) |
| M → S | 0.6857 (0.461, 0.974) | -34.53% (-6.73%, -54.98%) |
| S → A | 1.0737 (0.965, 1.19) | 56.29% (9.66%, 127.21%) |
| A → | 1.1009 (0.941, 1.278) | 2.82% (-12.8%, 22.0%) |
| **Charleston** |  |  |
| Before policy C | 2.4407 (1.73, 3.33) |  |
| C → N | 1.1092 (0.987, 1.24) | -54.63% (-37.5%, -67.2%) |
| N → R | 0.9840 (0.873, 1.104) | -11.28% (-11.0%, -11.6%) |
| R → A | 1.2526 (1.22, 1.28) | 27.20% (12.3%, 42.8%) |
| A → M | 0.8774 (0.855, 0.9) | -29.95% (-29.9%, -30.0%) |
| M → S | 0.9621 (0.927, 0.998) | 9.68% (8.48%, 10.93%) |
| S → | 1.0878 (1.07, 1.11) | 13.03% (10.9%, 15.1%) |
| **Colleton** |  |  |
| Before policy C | N/A |  |
| C → N | 1.3111 (0.75, 2.1) |  |
| N → R | 1.2956 (1.04, 1.59) | -0.39% (-41.5%, 76.4%) |
| R → M | 1.0228 (0.95, 1.10) | -21.20% (-2.42%, -35.47%) |
| M → S | 0.8782 (0.74, 1.03) | -13.97% (-27.06%, 0.89%) |
| S → A | 1.0353 (0.943, 1.133) | 17.78% (-1.33%, 38.12%) |
| A → | 1.1287 (1.05, 1.21) | 9.0% (-0.72%, 19.01%) |
| **Georgetown** |  |  |
| Before policy C | N/A |  |
| C → N | 1.3747 (0.938, 1.93) |  |
| N → R | 0.9440 (0.689, 1.256) | -31.32% (-49.23%, -8.12%) |
| R → A | 1.1980 (1.11, 1.29) | 26.38% (-5.19%, 69.56%) |
| A → M | 0.9596 (0.891, 1.032) | -19.90% (-12.8%, -26.5%) |
| M → S | 0.8468 (0.762, 0.937) | -11.70% (-1.96%, -20.34%) |
| S→ | 1.1016 (1.06, 1.14) | 29.90% (15.6%, 45.1%) |
| **Oconee** |  |  |
| Before policy C | N/A |  |
| C→ N | 1.2685 (0.674, 3.497) |  |
| N→ R | 1.0058 (0.718, 2.06) | -20.10% (-58.8%, 60.5%) |
| R→ M | 1.0645 (0.992, 1.362) | 5.19% (-21.9%, 44.3%) |
| M→ A | 1.0591 (0.901, 1.235) | -0.31% (-14.8%, 16.0%) |
| A→ S | 1.0259 (0.89, 1.17) | -3.16% (-1.21%, -4.91%) |
| S→ | 1.0710 (1.04, 1.1) | 4.25% (-9.85%, 20.29%) |
| **Orangeburg** |  |  |
| Before policy C | 1.7667 (0.745, 3.459) |  |
| C→ N | 1.0927 (0.746, 1.534) | -38.22% (-71.0%, 49.6%) |
| N→ R | 1.1872 (1.0, 1.39) | 8.24% (-22.3%, 52.5%) |
| R→ A | 1.2002 (1.13, 1.28) | 0.86% (-13.8%, 18.7%) |
| A→ M | 0.9585 (0.908, 1.011) | -20.16% (-13.5%, -26.7%) |
| M→ S | 0.8447 (0.763, 0.932) | -11.86% (-0.553%, -21.27%) |
| S→ | 1.0972 (1.06, 1.14) | 29.67% (16.0%, 44.1%) |
| **Richland** |  |  |
| Before policy C | 1.7828 (1.31, 2.36) |  |
| C→ N | 1.3423 (1.22, 1.47) | -24.77% (-0.05%, -43.81%) |
| N→ R | 0.9701 (0.911, 1.031) | -27.73% (-19.2%, -35.8%) |
| R→ A | 1.1729 (1.14, 1.21) | 20.86% (15.3%, 26.0%) |
| A→ M | 0.9529 (0.922, 0.984) | -18.76% (-18.7%, -18.8%) |
| M→ S | 1.0434 (1.01, 1.08) | 9.49% (9.39%, 9.6%) |
| S→ | 1.0439 (1.03, 1.06) | 0.016% (-1.78%, 1.79%) |
| **Williamsburg** |  |  |
| Before policy C | N/A |  |
| C→ N | 1.2495 (0.715, 2.001) |  |
| N→ R | 1.0775 (0.941, 1.227) | -13.65% (-48.1%, 52.1%) |
| R→ A | 1.1342 (1.02, 1.26) | 5.21% (-2.99%, 13.39%) |
| A→ M | 1.0013 (0.912, 1.096) | -11.75% (-1.1%, -21.4%) |
| M→ S | 0.8692 (0.767, 0.98) | -13.14% (-1.43%, -23.29%) |
| S→ | 1.1264 (1.07, 1.19) | 29.54% (12.4%, 48.3%) |

CrI, credible intervals. Labels – C: Closure of schools, N: Closure of non-essential businesses, R: reopening of nonessential businesses, A: County level mask mandate, M: state mask mandate, S: start of school, based on earliest date in the county. \*N/A indicates there is no reported case before Policy C (school closure) in the data after the 9-day shift to approximate the date of infection.

## Supplemental Figure Captions and Legends

**Supplemental Figure 1:**
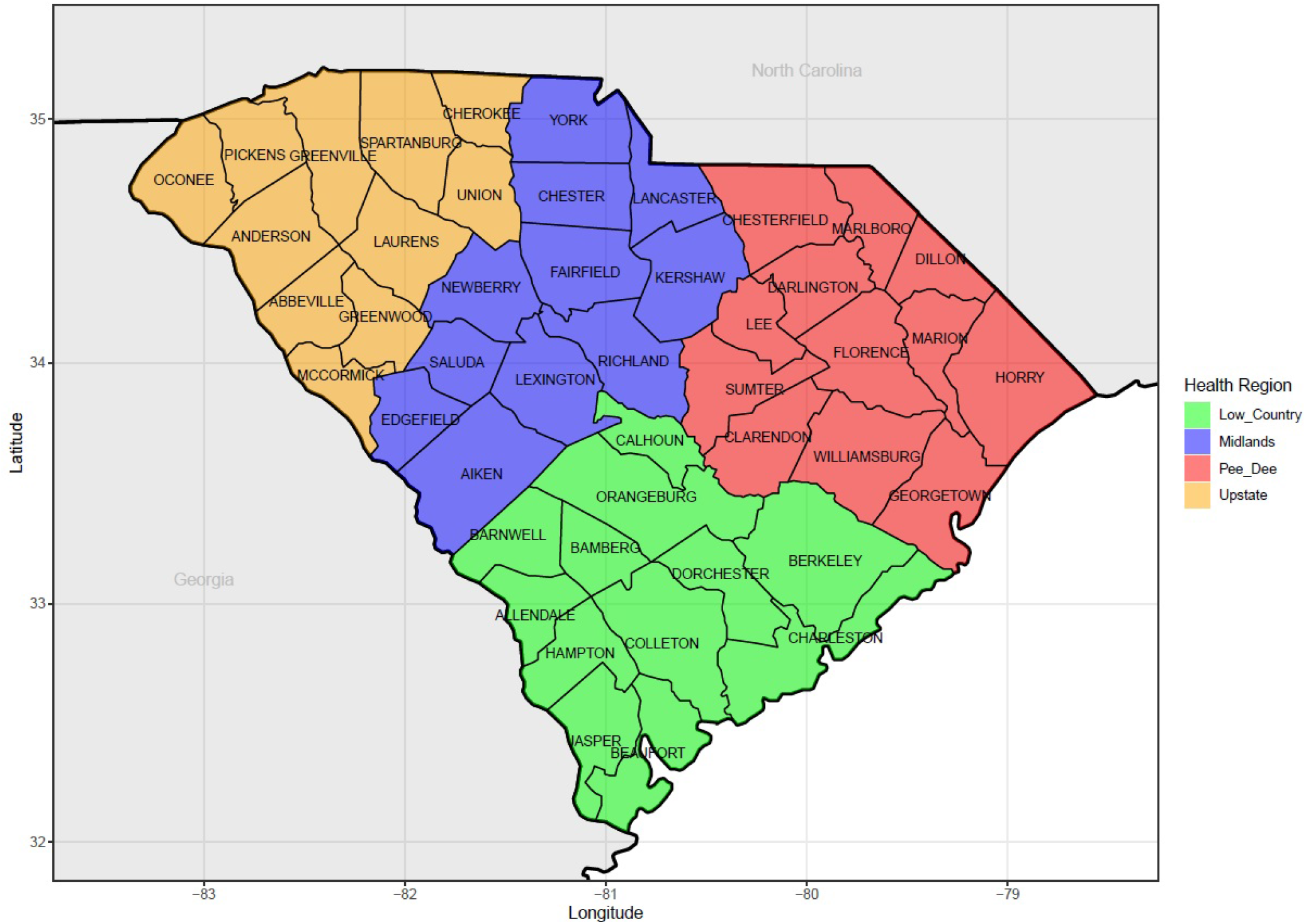
South Carolina Department of Health and Environmental Control (DHEC) region map. Regions are shown by color. Counties are labeled by name.

**Supplemental Figure 2.**
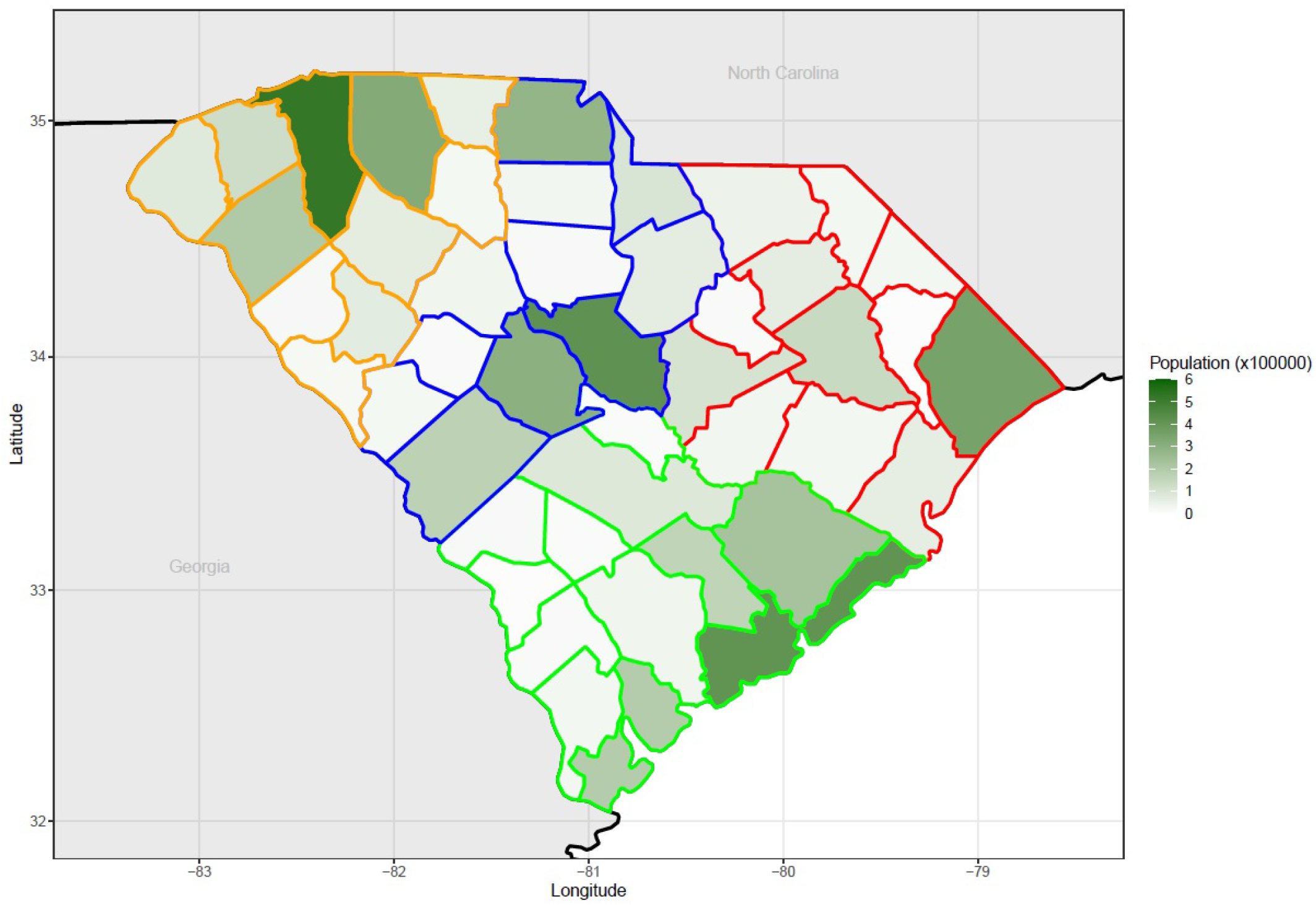
Population map of South Carolina. The Department of Health and Environmental Control (DHEC) health regions are outlined by color as shown in the previous figure.

**Supplemental Figure 3:**
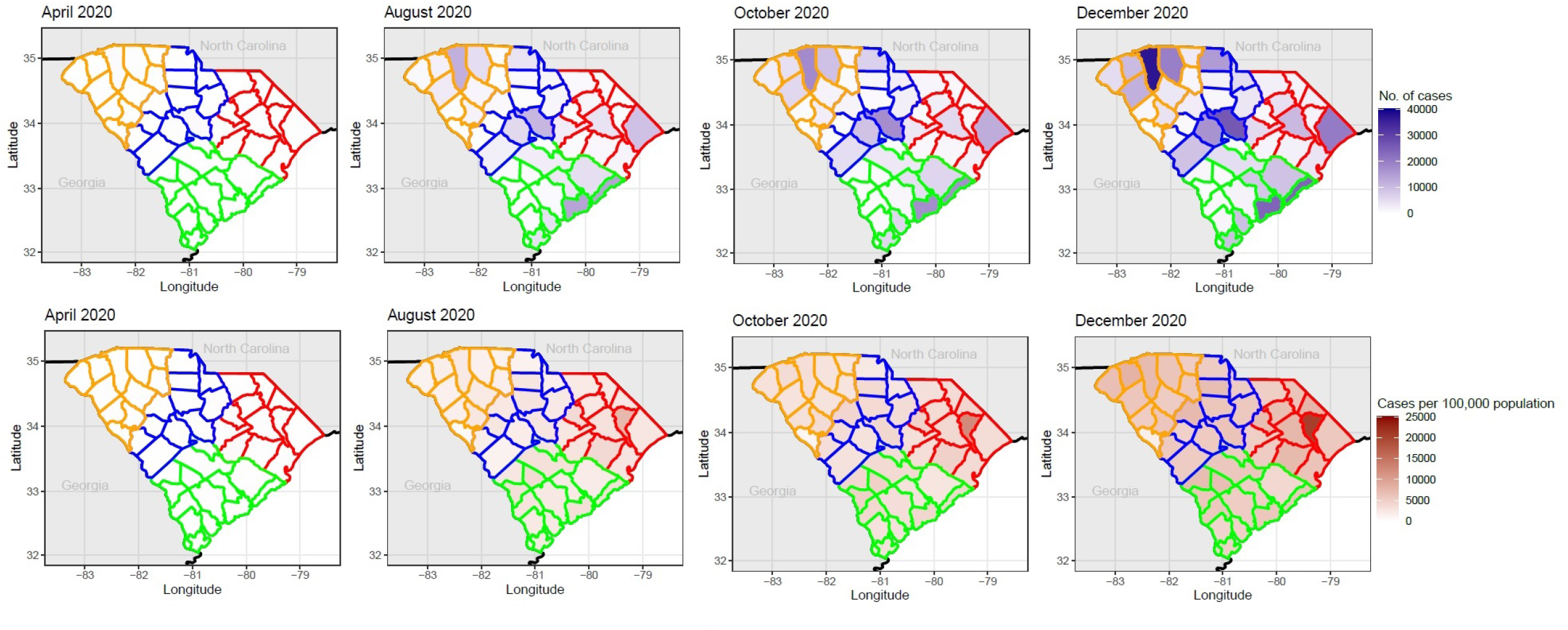
Maps of South Carolina counties group in Public Health Districts (county line color: Low Country: Green; Midlands: Blue; Pee Dee: Red; Upstate: Yellow) by cumulative case count (top 4 maps), and cumulative case counts per 100,000 population (bottom 4 maps) in April, August, October, and December, 2020.

**Supplemental Figure 4:**
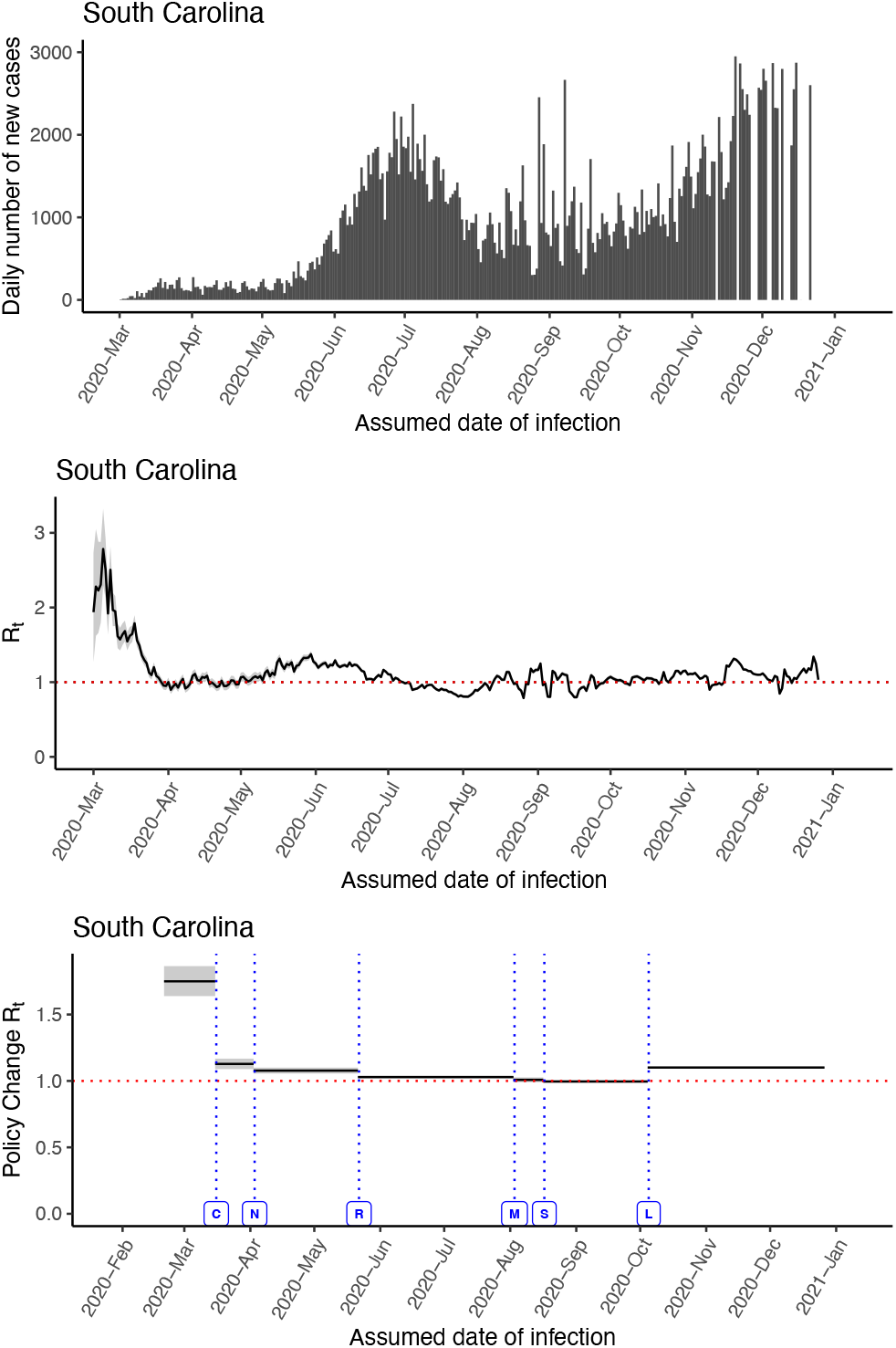
Sensitivity analysis: The incidence data is shifted backward by 15 days to approximate the date of infection, assuming a combined 15-day time lag of the incubation period and delay in testing.

**Supplemental Figure 5:**
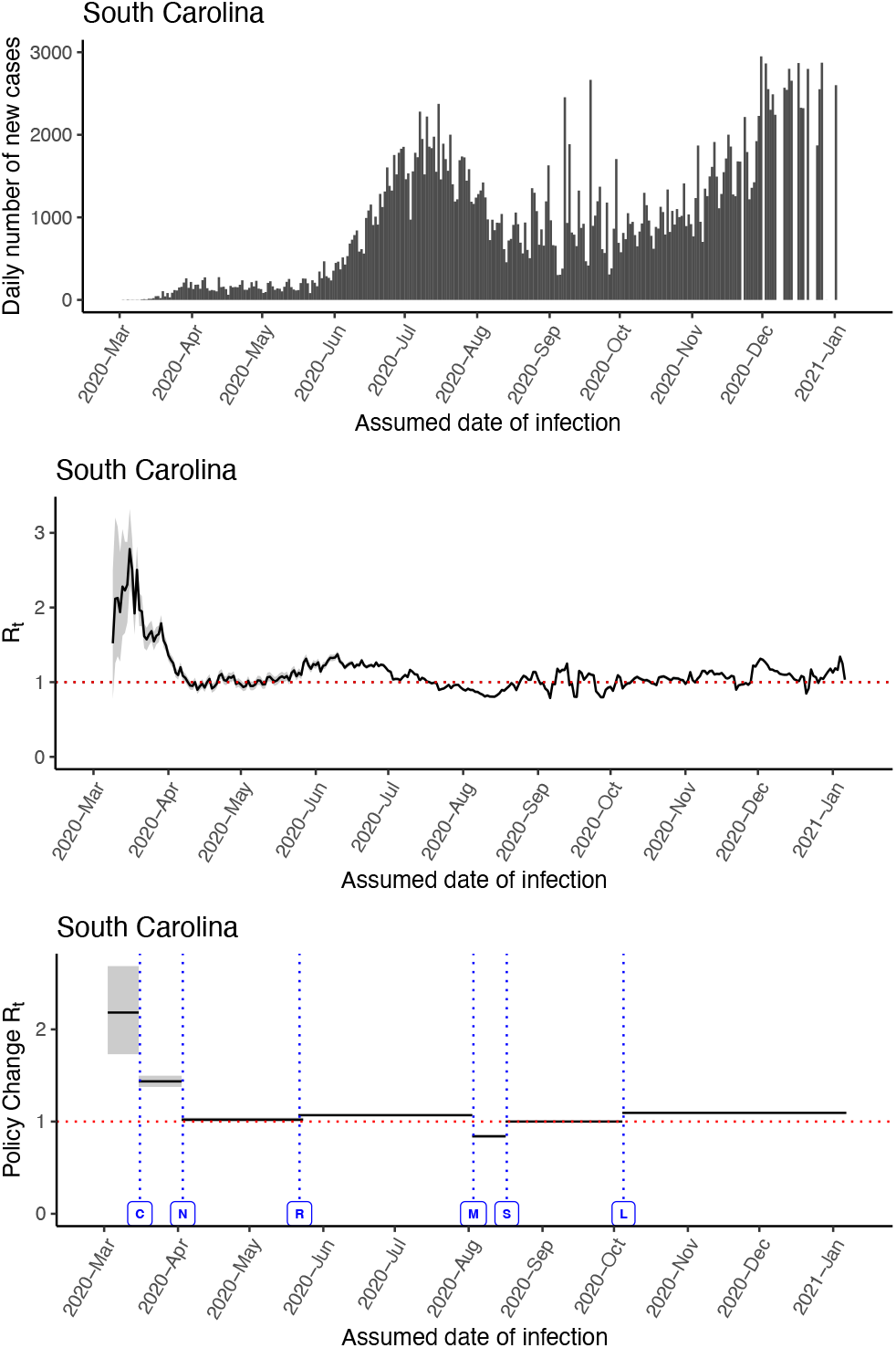
Sensitivity analysis: The incidence data is shifted backward by 4 days to approximate the date of infection, assuming a combined 4-day time lag of the incubation period and delay in testing.

